# Fibulin-3 as a reliable biomarker of fibrosis in obese subjects with Metabolically Dysfunction Associated Steatotic Liver Disease

**DOI:** 10.64898/2026.01.08.26343687

**Authors:** Allen Anthony Laraño, Silvia Palmisano, Deborah Bonazza, Discipio Marina, Marica Meroni, Anna Ludovica Fracanzani, Lory S Crocè, Claudio Tiribelli, Paola Dongiovanni, Natalia Rosso, Giraudi Pablo

## Abstract

**Introduction and Objectives:** Metabolic dysfunction-associated steatotic liver disease (MASLD) affects about one-quarter of adults worldwide, and liver fibrosis is its strongest predictor of liver-related morbidity and mortality. Using combined *in-silico* screening and clinical validation, we aimed to identify circulating biomarkers associated with fibrosis progression. Fibulin-3 was identified, and its diagnostic performance was evaluated in biopsy-proven MASLD cohorts.

**Materials and Methods:** The GSE125251 RNA-seq dataset was reanalyzed to compare liver transcriptomes from MASLD subjects with minimal (F0–F1) versus moderate to advanced fibrosis (F2/F3–F4). Differentially expressed genes (DEGs) were filtered to retain plasma-secreted, protein-coding candidates. Top-ranked genes were evaluated in liver biopsies from a morbidly obese cohort (n = 65) stratified by fibrosis stage, and their plasma levels were measured via ELISA in an independent bariatric cohort (n = 225).

**Results:** Among 106 DEGs, 22 encoded plasma-circulating proteins. Six top candidates (EFEMP1, LTBP2, LUM, DPT, CHI3L1, CCL20) were prioritized. EFEMP1 (Fibulin-3) showed the strongest association with fibrosis, with significantly higher hepatic mRNA and protein expression in F2/F3–F4 versus F0–F1 (p < 0.005). Plasma Fibulin-3 levels correlated with fibrosis stage (ρ = 0.40, p < 0.0001), increasing from 9.4 ng/mL in F0–F1 to 21.7 ng/mL in F2/F3–F4. Its diagnostic performance for F ≥ 2 (AUROC = 0.78) exceeded that of APRI, FIB-4, NFS, and HSI. A combined index including Fibulin-3, HSI, platelets, and GGT increased the AUROC to 0.87 (CI: 0.79–0.92).

**Conclusions:** Plasma Fibulin-3 is notably higher in individuals with advanced MASLD and represents a promising non-invasive biomarker for liver fibrosis stratification in metabolically unhealthy obese populations.

## 1. Introduction

Metabolic dysfunction-associated steatotic liver disease (MASLD) is characterized as a spectrum of liver diseases caused by the excessive accumulation of fat in hepatocytes (1–4) . This condition is particularly associated with high-risk metabolic syndromes such as overweight, hypertension, type 2 diabetes mellitus, insulin resistance, and dyslipidemia, while excluding secondary causes (e.g., drug toxicity, alcohol consumption, and viral infection) (5). The global prevalence of MASLD among adults is estimated to be 25.2% (range 22.1%-28.6%). It is predicted to increase due to the increasing incidence of obesity and T2DM among adults and children (2).

MASLD encompasses a spectrum of hepatic pathological manifestations, ranging from simple steatosis (excessive liver fat) to metabolic dysfunction-associated steatohepatitis (MASH), characterized by hepatocellular injury, inflammation, and varying degrees of liver fibrosis. This condition can ultimately progress to cirrhosis and, ultimately, hepatocellular carcinoma (3–7). The progression of MASLD is a continuum, with liver fibrosis representing the most critical histological predictor of mortality in MASLD patients (6,7). Therefore, the early identification of obese and non-obese subjects with liver fibrosis is essential to mitigate its development and improve prognosis (8).

The current gold standard for diagnosing MASLD and staging its progression remains liver biopsy. However, this procedure is costly, invasive, and carries the risk of complications such as bleeding and trauma. As a result, identifying reliable biomarkers represents a promising alternative for developing non-invasive, rapid, and accurate tools to distinguish MASH and fibrosis within the MASLD spectrum. In recent years, several studies employing multi-omics approaches -including transcriptomics, metabolomics, and proteomics- have been conducted to individuate potential biomarkers for MASH and fibrosis in MASLD (9–12). As an example, previous research has highlighted various inflammatory markers associated with MASH diagnosis, including C-reactive protein (*CRP*), tumor necrosis factor (*TNF*), interleukin-8 (*IL-8*), and C-X-C motif chemokine ligand 10 (*CXCL10*) (9,10). Also, biomarkers that indirectly reflect liver fibrosis have been reported, including tissue inhibitor of metalloproteinases 1 (TIMP1), procollagen type III N-terminal peptide (*PIIINP*), cytokeratin-18 (*CK18*), and apolipoprotein A-IV (*ApoA-IV*) (11–14). Furthermore, numerous non-invasive algorithms have been developed to assist in fibrosis staging for patients with MASLD, including the Forns score, Fibrosis - NASH Index (FNI), and the MACK-3 score, which integrate various clinical and biochemical parameters (15, 16). The Fibrosis-4 index is utilized as a first-line screening tool for liver fibrosis (13). Although several candidate biomarkers and predictive models for fibrosis diagnosis in MASLD have been identified, their validation and application in routine clinical decision-making are still in early stages.

In this study, we utilized publicly available transcriptome datasets and developed an *in-silico* biomarker discovery strategy. Several advantages support this method: transcriptome profiles are among the most accessible omics datasets in MASLD research; these data are typically normalized and harmonized; and numerous bioinformatics tools are available to facilitate data integration through meta-analysis (15,16). Our investigation identified biomarkers for evaluating MASH-induced fibrosis using this *in-silico* approach. Among the candidate biomarkers, Fibulin-3 was selected for further study to assess its diagnostic ability to distinguish fibrosis in MASLD, particularly within a high-risk group of morbidly obese (MO) patients.

## 2. Patients and methods

### 2.1 Discovery of biomarkers through an in-silico identification approach

The in-silico strategy was initiated by retrieving bulk-transcriptome datasets from the NCBI-GEO database (112). The GSE13525 dataset, containing RNAseq data of 206 individuals with MASLD, was recategorized according to liver histology data into two groups: 139 individuals with early fibrosis stage (F0-F1) and 67 individuals with moderate/advanced fibrosis stage (F2, F3, F4) (Kleiner-Brunt Fibrosis scoring system). The 3D-RNA Seq App pipeline was used to determine differential gene expression profiles between these two groups (14). To select the candidate genes, we were interested in genes encoding secreted proteins; the secretome and plasma proteome data were retrieved from the Human Protein Atlas (HPA) and the Plasma Proteome Databases, respectively (15,16). Venn diagrams were used to identify candidate biomarkers from the pool of differentially expressed genes. Those fulfilling the following desired criteria were selected: genes coding for secreted proteins, released in blood, and the exclusion of housekeeping proteins was considered (17).

Gene Pathway Enrichment Analysis was conducted using the gProfiler platform to visualize the functional profiles of key genes and gene sets based on information from Gene Ontologies (GO), Reactome (REACT), and the Kyoto Encyclopedia of Genes and Genomes (KEGG) databases. GO terms for Molecular Function (GO: MF), Biological Process (GO: BP), and Cellular Component (GO: CC), along with REACT and KEGG terms with P < 0.05, were considered statistically significant and included in the enrichment plots (18,19).

### 2.2 Study Design and Participants

This retrospective study involved experimental determinations in samples from two healthcare institutions that perform bariatric surgeries, located in Trieste and Milan. Overall, samples and clinical data from 225 subjects (157 female, 68 males, ages 44.8 ± 9.6) with severe obesity who consecutively underwent bariatric surgery according to international criteria [i.e. body mass index (BMI) >40 or >35 when a major comorbidity (i.e. diabetes, obstructive sleep apnea syndrome) co-existed] were included in the study. The only exclusion criteria were HBsAg and/or anti-HCV positivity and an intake of more than 30 or 20 gr/die of alcohol in male or female patients.

One hundred ten obese subjects consecutively enrolled underwent bariatric surgery at Trieste University Hospital, while 115 obese subjects at Fondazione IRCCS Cà Granda, Ospedale Maggiore Policlinico of Milan. These groups were included as cohorts for the discovery and validation study phases, respectively. All patients provided written consent, and anonymity and protection of sensitive data were maintained through pseudonymization of all collected samples for the research. The respective local Ethical Committees approved the studies. Two pathologists, blinded to clinical data, interpreted the liver specimens collected during surgery (wedge biopsy). Steatosis was graded based on fat content (lipid droplets) in hepatocytes, as seen in hematoxylin and eosin-stained samples. Biopsies showing no or minimal (<5%) steatosis and no injury or fibrosis were classified as normal. Samples with more than 5% steatosis were labeled as MASLD. The histological diagnosis of MASH and fibrosis was made according to Kleiner-Brunt criteria.

Sixty overweight or obese subjects consecutively enrolled at the Liver Disease Center (Centro Patologie del Fegato, Ospedale Maggiore, Trieste, Italy) were included as a secondary validation cohort. All participants underwent non-invasive imaging for fibrosis assessment using point Shear Wave Elastography (pSWE) with the Elasto2D protocol on a Philips Epiq ultrasound system (Philips Healthcare, Best, The Netherlands) equipped with a 1–5 MHz convex probe. Liver stiffness was quantified from the velocity of shear wave propagation, expressed in kilopascals (kPa), as a surrogate of parenchymal elasticity. Significant fibrosis was defined according to validated pSWE cut-off values: F0–F1: <5.7 kPa; F2: 5.7–7.99 kPa; F3–F4: >8 kPa (20).

### 2.3 Plasma biomarkers’ quantification

Plasma protein levels were measured using the following commercial ELISA kits: Fibulin-3 (Human FBLN3 ELISA Kit, E-EL-H1673, Elabscience), Lumican (Human Lumican ELISA Kit, E1871Hu, BT Lab), and Chitinase 1-like 3 (RayBio® Human CHI3L1 ELISA Kit, ELH-CHI3L1, RayBiotech) following the manufacturers’ instructions. For additional information about methods, such as molecular techniques like RNA extraction, relative gene expression quantification by qPCR, protein analysis by Western blot, and further details on ELISA determinations, see the Supplementary Information.

### 2.4 Statistical Analysis

The MO cohort (n=225) was divided, as mentioned earlier, into two subsequent cohorts: the discovery MO cohort, which included 110 MO subjects and adjusted the fibrosis prevalence to 37% (enrichment of the moderate/advanced fibrosis proportion), and the validation MO cohort, which included 115 MO subjects and maintained the fibrosis prevalence (33%), close to that of the global MO population (24%)(21). In both the MO discovery and validation cohorts, subjects were stratified according to fibrosis stage (F0-F1, minimal fibrosis; F2/F3-F4, moderate/advanced fibrosis) to assess the best candidate based on diagnostic performance analysis (accuracy and determination of optimal cut-off values). Continuous variables were expressed as mean ± standard deviation and categorical as numbers or percentages. Categorical variables were analyzed using chi-square tests with correction, when appropriate. Independent t-test and ANOVA were used for normally distributed continuous variables. Non-parametric tests (Mann–Whitney, and Kruskal–Wallis) were applied for ordinal or continuous variables that failed to pass D’Agostino & Pearson omnibus normality test. Correlation analyses were performed using Spearman’s correlation coefficients to estimate the correlation of plasma levels and several factors of interest. Statistical analysis was performed by using GraphPad Prism version 10.01 software. Logistic regression analysis was used to identify independent factors associated with fibrosis. The predictive model was constructed, including the four most strongly associated variables (independent factors) selected after applying the hierarchical forward selection algorithm in the subset selection test mode performed with NCSS statistical software (version 12.0.16). The diagnostic performance of the selected candidate, Fibulin-3, was assessed using receiver operating characteristic (ROC) curves. The area under the ROC (AUROC) using empirical method was used to compare the accuracy among the different fibrosis diagnostic tests. The sensitivity, specificity, positive predictive values (PPVs), and negative predictive values (NPVs) for relevant cut-offs were also calculated. ROC analyses were performed using NCSS statistical software (version 12.0.16). Statistical significance was determined at p < 0.05. All figures and statistical analyses were generated using GraphPad Prism 10 and NCSS statistical software (version 12.0.16), respectively.

## 3. Results

### 3.1 In-silico identification of candidate biomarkers involved in MASH-fibrosis

Identifying candidate protein-coding genes linked to liver fibrosis began with a differential expression analysis between moderate/advanced (F2-F3-F4) and minimal fibrosis (F0-F1) cohort groups. 106 differentially expressed genes were identified; 16 genes were downregulated, while 90 were upregulated in the moderate/advanced group (**Figure 1A**). The complete list of DGEs is mentioned in the Supplementary Table 1 **(Table S1)**. Subsequently, to determine the molecular pathways that were overrepresented by these differentially expressed genes, a functional enrichment analysis was conducted using gProfiler. **Figure 1B** shows the top GO, KEGG, and Reactome pathway terms, indicating that the DGE candidates are primarily involved in extracellular matrix (ECM) organization, chemokine activity, and elastic fiber formation, all of which are biological processes related to fibrogenesis during liver tissue injury.

**Figure 1.**
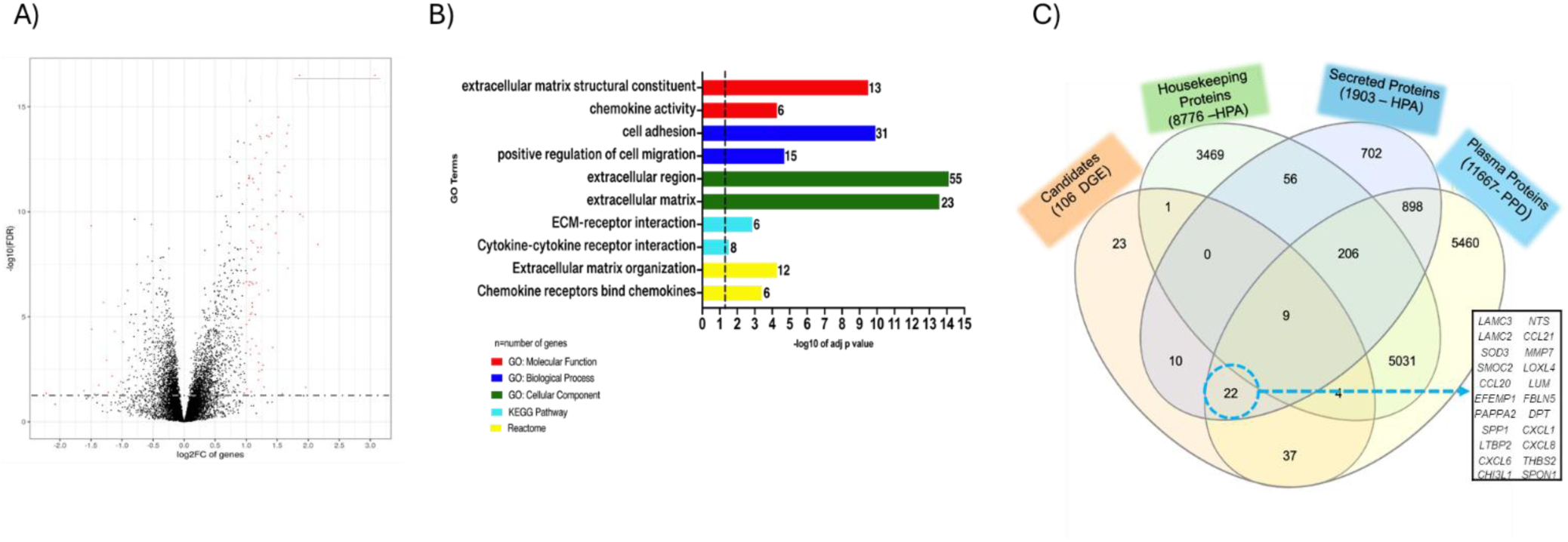
Differential expressed genes using 3D-RNAseq pipeline on publicly available GSE135251 dataset. (A) Volcano plot showing DE genes: log2FC vs -log10(FDR) at gene level. The red dots represent the significantly dysregulated genes. B). Top GO terms and pathways enriched by Gene Ontology, Kyoto Encyclopedia of Genes and Genomes and Reactome based on Functional Enrichment Analysis of upregulated genes using gProfiler. The dashed line is the threshold line for statistically significant terms using Fisher’s T-test. C)Venn diagram illustrating the number of secretory plasma proteins identified as differentially expressed by pairwise analyses using MASL or MASH F0/F1 as a baseline. Housekeeping and secreted protein datasets were obtained from the Human Protein Atlas (HPA) and the Plasma protein dataset from the Plasma Proteome Database (PPD).

Additionally, the 106 differentially expressed genes were analyzed alongside various datasets from the Human Protein Atlas and the Plasma Proteome Database using a similar *in-silico* approach, as fully described by Giraudi et al. (41, 79). After dataset integration, candidate secretory plasma proteins were filtered using Venn diagrams, and those meeting the selection criteria were selected from the total DGEs (**Figure 1C**). The candidate protein-coding genes proposed as potential biomarkers are listed in **Table 1**. Among the comprehensive list of identified candidates, eight protein-coding genes relevant to fibrogenic signaling and ECM remodeling were subjected to experimental testing.

**Table 1.**
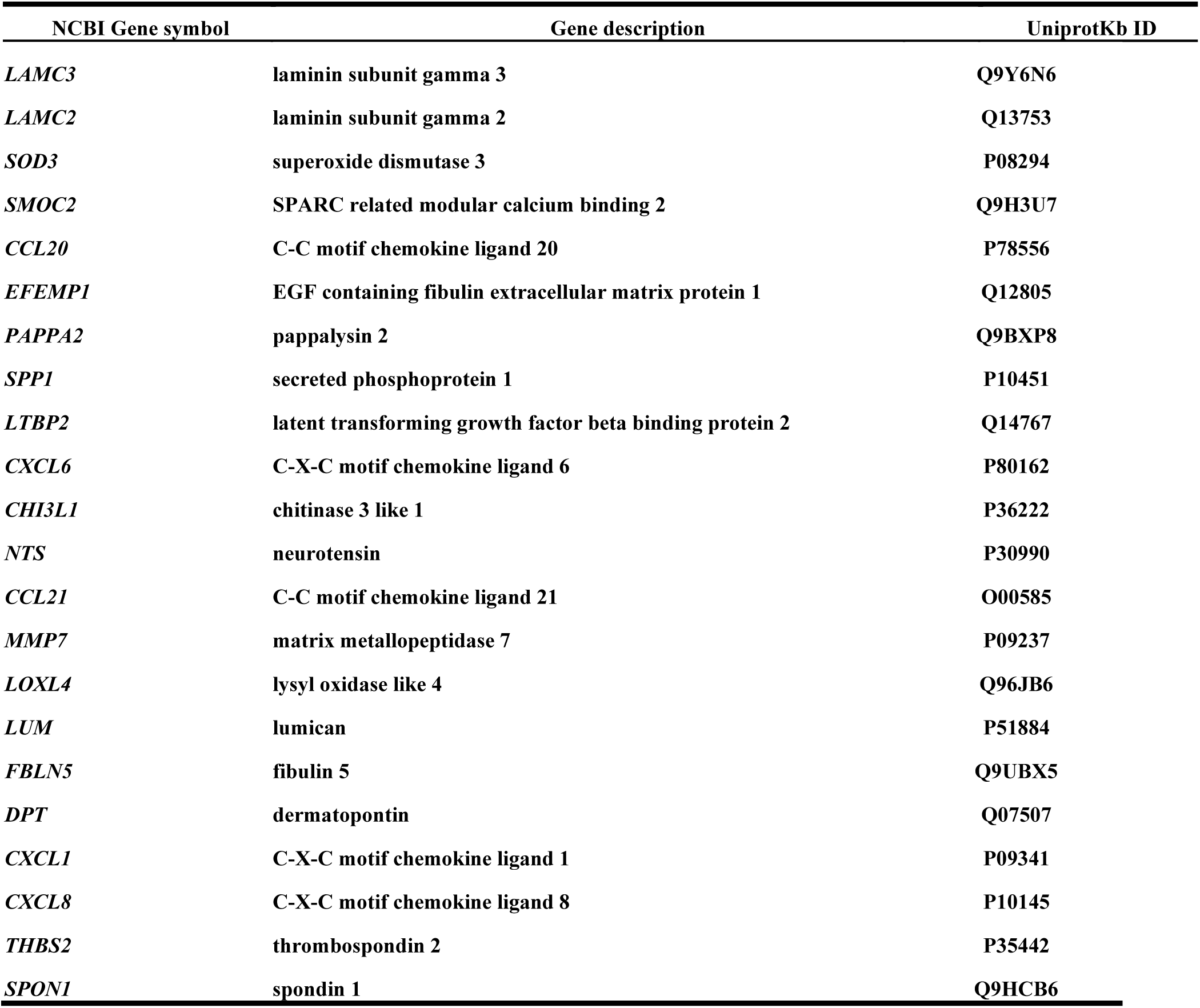
Candidate biomarkers identified by the *in-silico* strategy.

The clinical demographic data for the patients in the Trieste discovery cohort, Milan validation cohort, and combined MO cohort (n=225) are reported in **Table 2**. Statistics showed significant differences for age (p < 0.003), with younger subjects having reduced total cholesterol (p = 0.01) and presenting a higher liver steatosis grade (p < 0.001) in the Milan validation cohort. This data shows that both cohorts are comparable for the rest of the clinical and histological parameters.

**Table 2.**
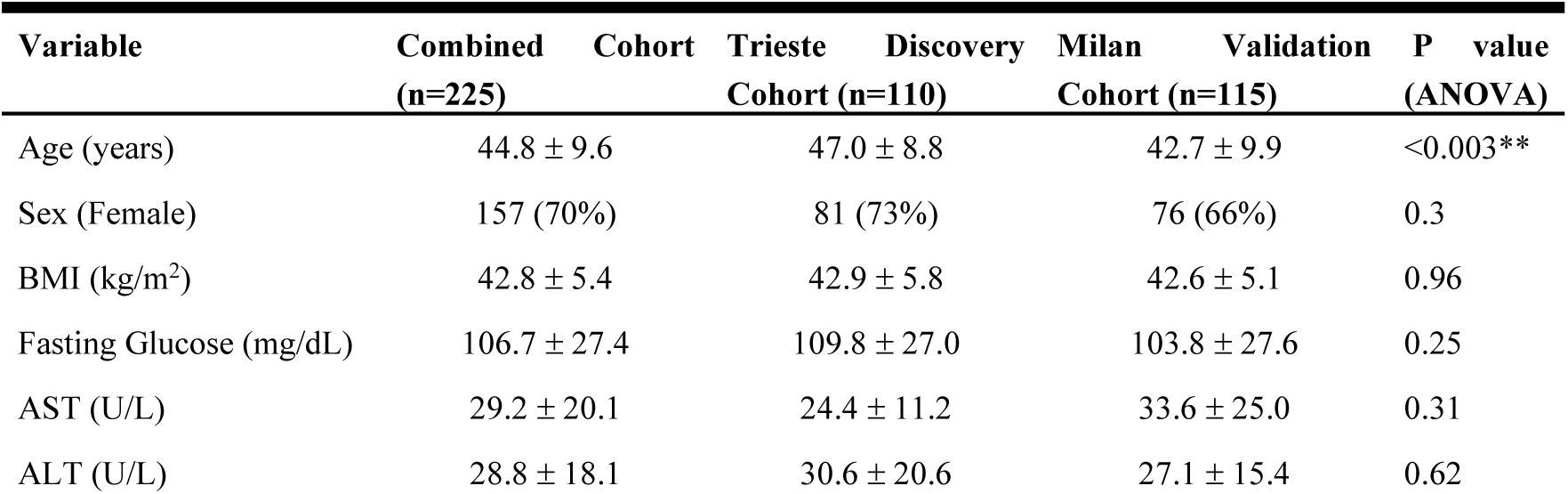
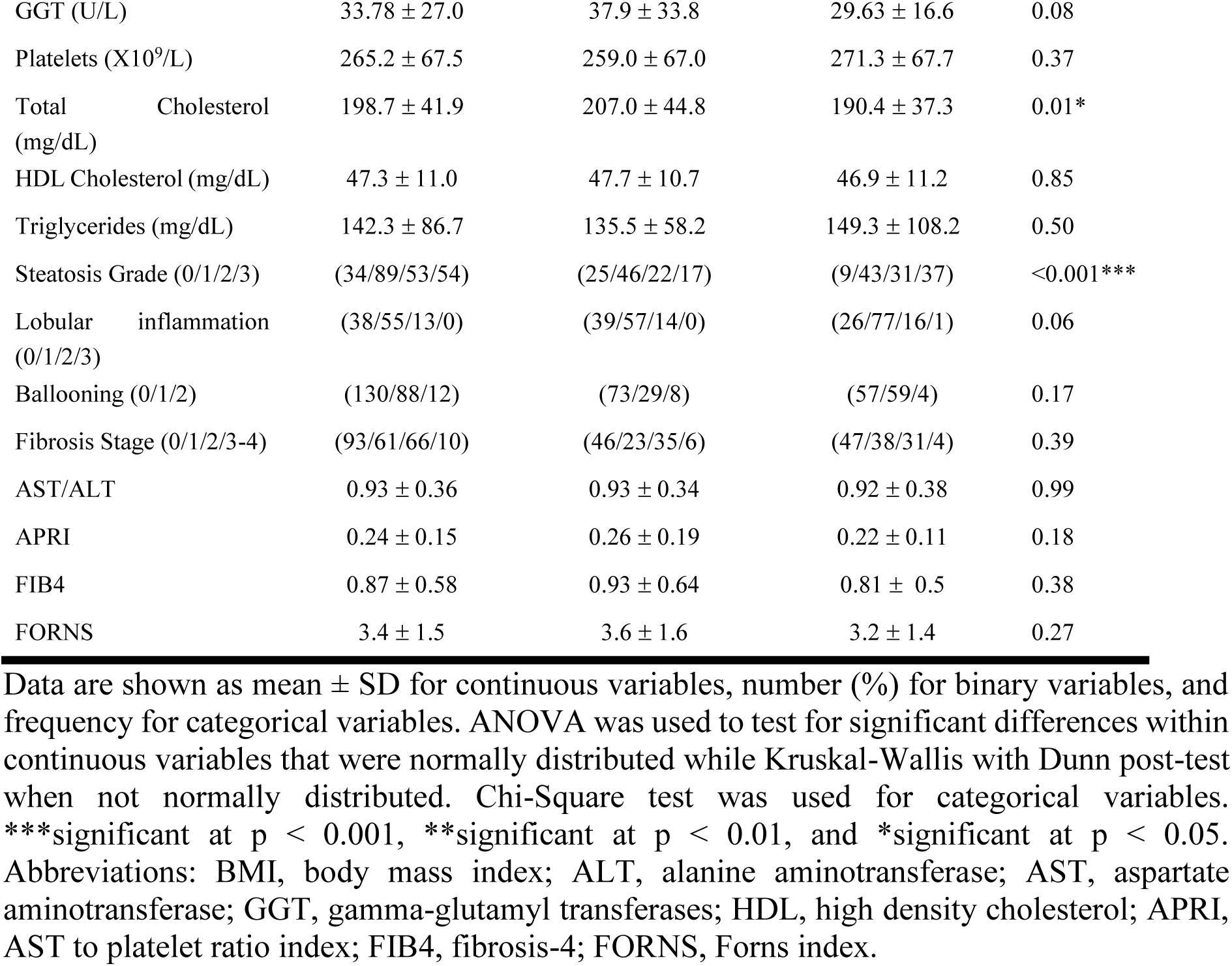
The clinical-demographic data of the patients selected in the discovery, validation, and combined MO cohorts.

A subset from the MO Trieste discovery cohort of 65 liver biopsies, categorized into F0-F1 and F2-F3-F4 fibrosis groups (see **Table 3**), was used to verify changes in liver gene expression of the eight candidate markers. Based on the mRNA expression analysis, *EFEMP1, CHI3L1, and LUM* exhibited the most significant changes from early to moderate fibrosis (p = <0.0001). Specifically, *EFEMP1* mRNA expression in the liver increased 2.2-fold (p < 0.001), *CHI3L1* increased 5.5-fold (p = 0.006), and *LUM* increased 1.6-fold (p = 0.006) in moderate fibrosis compared to early fibrosis (**Figure 2A**).

**Figure 2.**
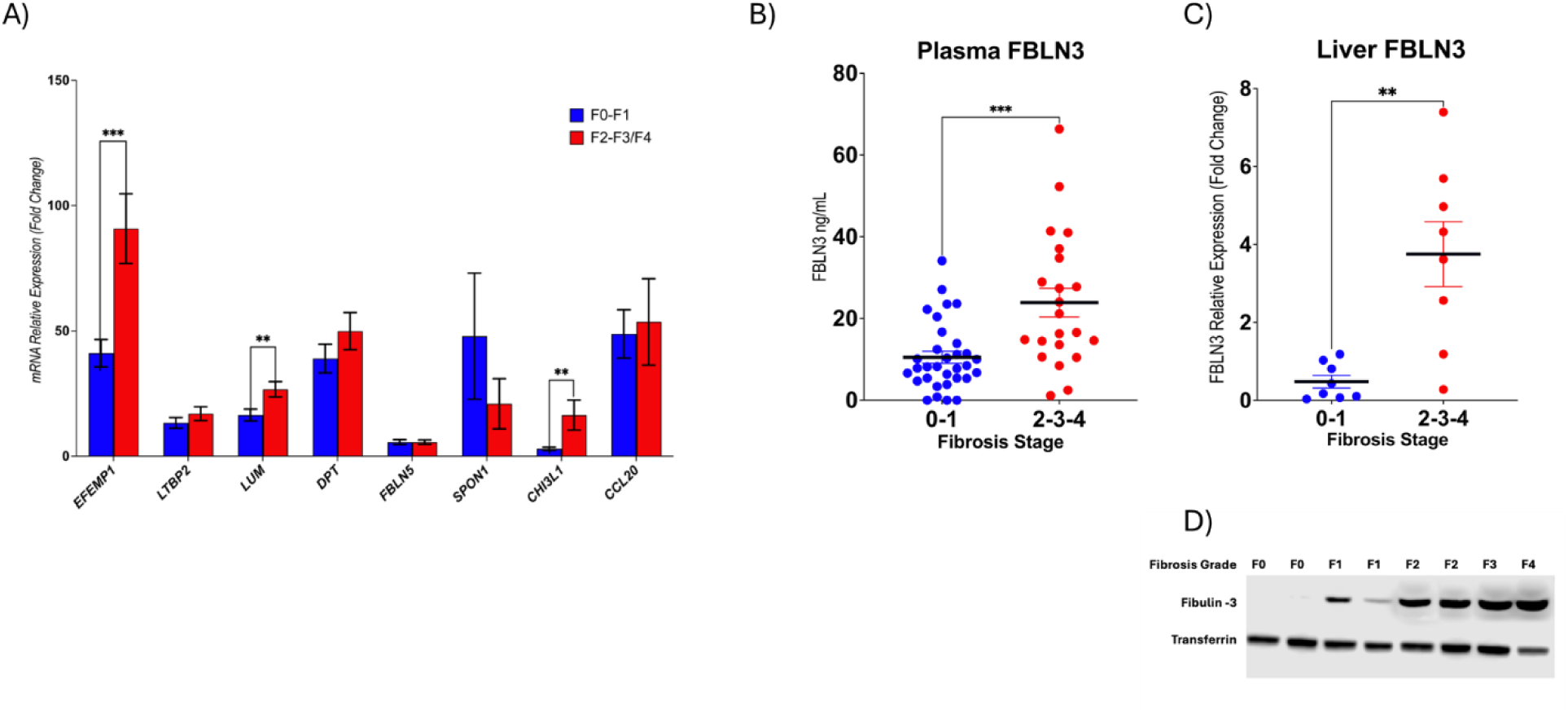
Gene and protein expression assays were in samples of the Trieste discovery cohort. A) mRNA relative expression analysis of 8 protein-coding genes candidates in liver samples (n=54), B) Fibulin-3 plasma levels by ELISA in the first analyzed batch of the Trieste discovery cohort (F0-F1, n = 30; F2-F3-F4, n = 30) and C) Representative Western blot showing Fibulin-3 protein levels in liver homogenates from subjects with different fibrosis stages. D) Western blot detection of Fibulin-3 (EFEMP1) in liver tissue homogenates. Bands corresponding to Fibulin-3 (∼55 kDa) and transferrin (∼80 kDa) are shown. Relative Fibulin-3 expression was quantified by densitometric analysis and normalized to transferrin to account for protein loading variations across samples. Values presented are the mean ± SD *p <0.05, **p <0.01, ***p <0.0001.

**Table 3.**
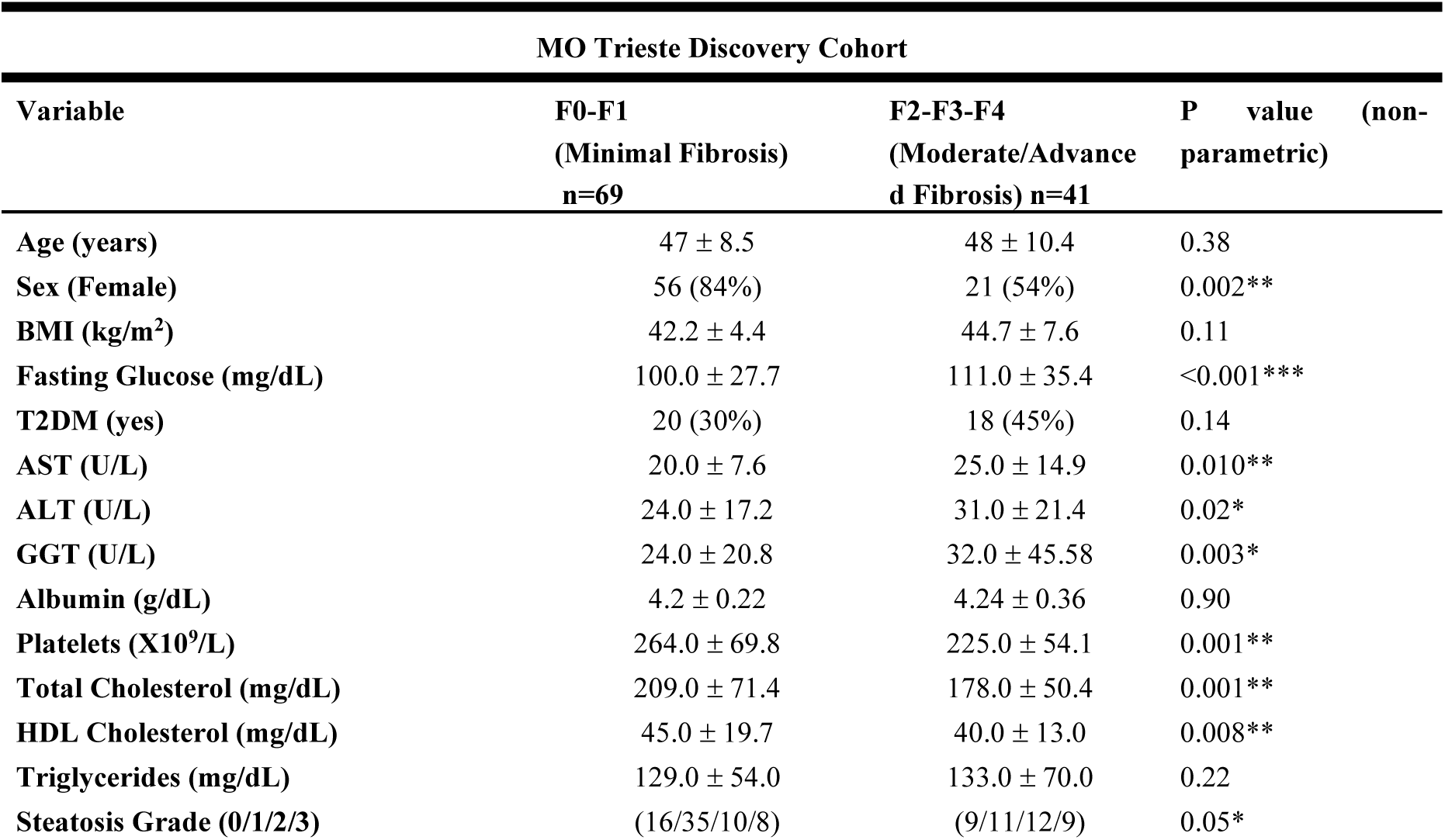
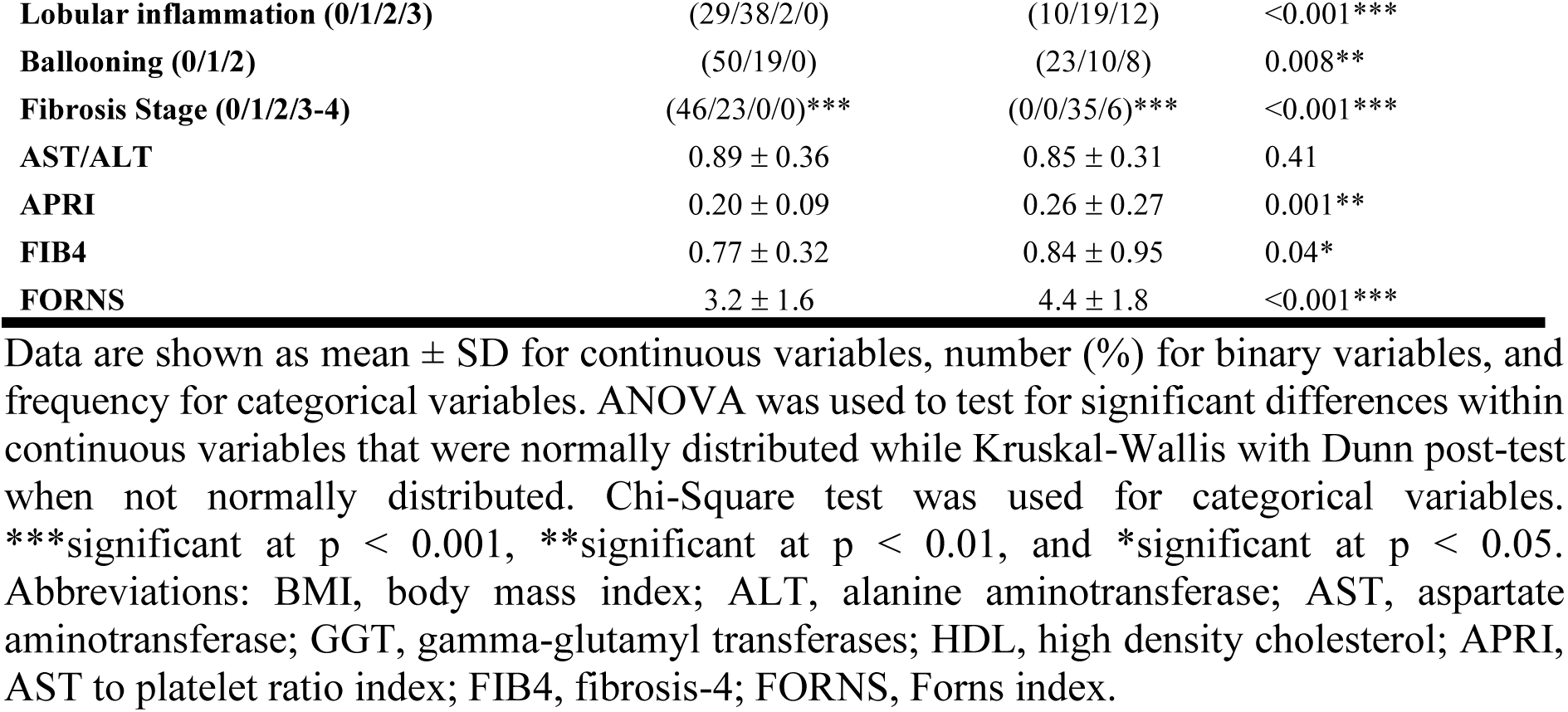
The clinical-demographic data of the patients selected in the discovery cohort stratified by fibrosis stage.

### 3.2 Liver and Plasma fibulin-3 protein levels are elevated in hepatic fibrosis

Next, we aimed to determine whether changes in liver gene expression were also reflected in circulating blood proteins. Therefore, we measured the plasma levels of the encoded protein products of *EFEMP1*, *Lum*, and *CHI3L1* genes (Fibulin-3, Lumican, and Chitinase 3-like 1 proteins, respectively) using ELISA kits. Fifty-four plasma samples from the Trieste discovery cohort were initially analyzed, and pairwise comparisons were made between the fibrotic groups. Our data showed that Fibulin-3 plasma levels were significantly higher in moderate/advanced fibrosis, increasing from 10.4 ± 8.4 ng/mL in F0-F1 to 23.9 ± 16.3 ng/mL in F2-F3-F4 samples (**Figure 2B**). In contrast, there were no significant differences in plasma levels of Lumican and Chitinase-3-like 1 proteins between the two groups (shown in **Supplementary Figure S1 A and S1 B**). Additionally, we examine Fibulin-3 protein expression by Western blot in a subset of 16 liver samples from the same cohort. As shown in **Figure 2C-2D**, hepatic Fibulin-3 levels increased by 7.8-fold (p=0.002) in F2–F3-F4 samples compared to F0–F1 samples, reflecting the trend observed in plasma levels.

### 3.3 Diagnostic performance of plasma fibulin-3 for liver fibrosis

Having established that Fibulin-3 is linked to liver fibrosis and that plasma changes reflect the observed alterations at the hepatic level, we assessed the diagnostic performance of plasma Fibulin-3 by increasing the number of plasma samples from MO patients.

First, plasma levels of Fibulin-3 were measured in 110 samples from the Trieste discovery cohort. **Table 3** presents clinical-demographic data for the patients included in the Trieste discovery cohort stratified into two fibrotic groups. Fibulin-3 levels effectively can distinguish fibrosis stages within the Trieste cohort, with F0–F1 samples presenting median levels of 10.7 ± 7.7 ng/mL and significantly higher concentrations observed in F2–F3/F4 samples, reaching 24.4 ± 17.1 ng/mL (**Figure 3A**). Additionally, Fibulin-3 plasma levels increased progressively with fibrosis severity in both males and females (p < 0.001), with no significant differences between sexes (p = 0.25) and no interaction between sex and fibrosis stage (p = 0.13), indicating that the association between plasma Fibulin-3 and fibrosis progression is independent of sex (**Figure 3B**). Furthermore, small differences in plasma Fibulin-3 levels were observed across different grades of steatosis (p=0.006) and lobular inflammation (p=0.02), but no significant difference was found in ballooning (p=0.16) (**Figure 3C-F**).

**Figure 3.**
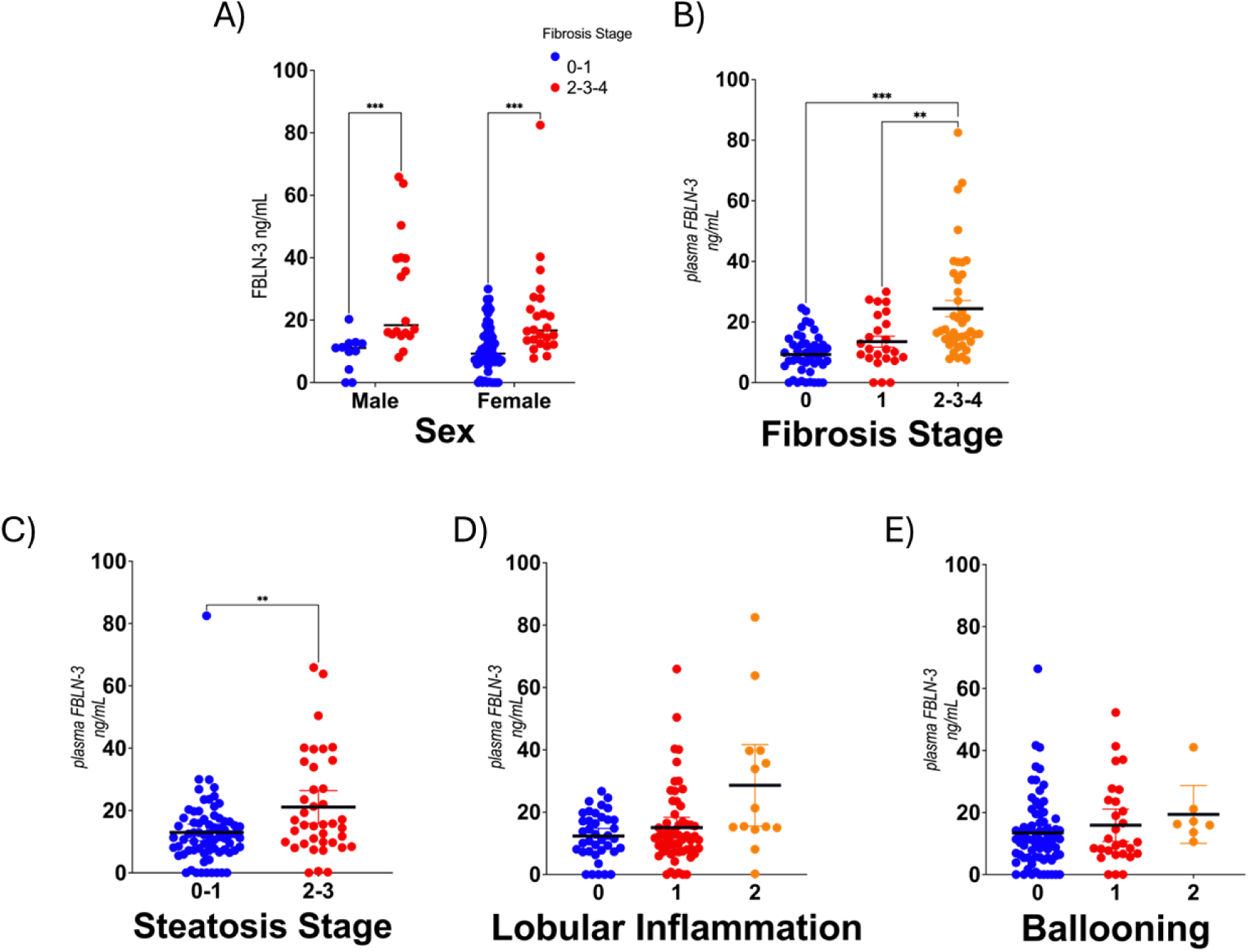
Comparison of plasma FBLN-3 levels across different parameters in the Trieste discovery cohort (n = 110) are shown as follows. A) Comparison between F0/F1 and F2-F3/F4 fibrosis stages across sex, B) Comparison across fibrosis stages, C) Comparison across steatosis stage, D) Comparison across lobular inflammation grade and E) Comparison across ballooning grade.

To assess the reproducibility of Fibulin-3 as a circulating biomarker, we examined its performance in an independent validation cohort, the Milan validation cohort (n = 115). Receiver operating characteristic (ROC) curve analysis demonstrated that, in the Trieste discovery cohort, plasma Fibulin-3 achieved an area under the ROC curve (AUROC) of 0.82 (IQR 0.73-0.89), with 78% sensitivity and 74% specificity at an optimal cut-off of ≤ 12.9 ng/mL (**Figure 4A**). A slightly lower AUROC of 0.73 (IQR 0.61-0.82), was observed in the Milan validation cohort, with 49% sensitivity and 90% specificity at a cut-off of ≤ 16.8 ng/mL (**Figure 4B**). When both cohorts were combined (n = 225), plasma Fibulin-3 maintained robust diagnostic performance with an AUROC of 0.78 (IQR, 0.71-0.81), achieving 65% sensitivity and 80% specificity at a cut-off of ≤ 14.8 ng/mL (**Figure 4C**). Notably, comparison of AUROCs using the empirical approach revealed that Fibulin-3 (AUROC 0.78) outperformed commonly used non-invasive fibrosis scores, including APRI (0.67), FIB-4 (0.64), FORNS (0.68), and NFS (0.50), as shown in **Figure 4D**.

**Figure 4.**
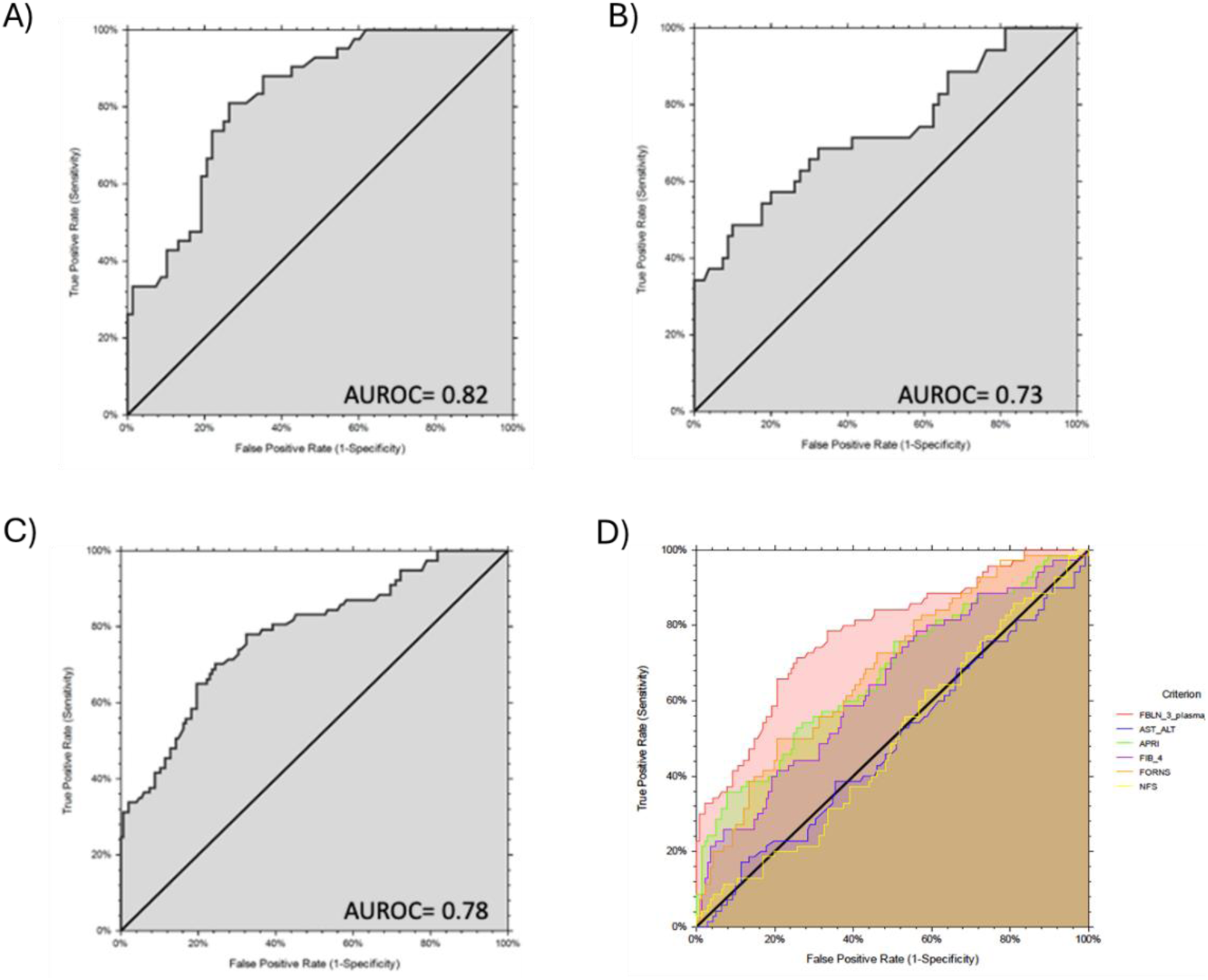
AUROC curves for plasma FBLN-3 are shown as follows. (A) from the discovery cohort, B) from the validation cohort, C) from the combined cohort and D) Comparison of AUROC curves of the plasma FBLN-3 against other blood-based scoring tests.

Additionally, we wanted to assess whether the Fibulin-3 marker might also be useful in overweight or obese subjects, so we evaluated the diagnostic performance in a second, independent cohort, the Centro Patologie del Fegato (CPF) cohort, which mainly includes overweight and obese individuals. **Table S2** provides the clinical-demographic data for patients in the CPF cohort, divided into two groups: 30 F0-F1 and 30 F2-F3-F4 plasma samples. In this cohort, the median plasma levels of Fibulin-3 were 8.9 ± 6.6 ng/mL in the F0-F1 group, with a slight increase in F2–F3-F4 samples, reaching a mean of 14.4 ± 8.1 ng/mL (p = 0.002) (Supplementary material **Figure S2**). Furthermore, an AUROC of 0.71 (IQR 0.578 to 0.820) was observed. Fibulin-3 AUROC also outperformed AST/ALT (0.56), APRI (0.53), and FIB-4 (0.62) (**Figure S2 B**).

To further analyze the MO cohorts (Trieste and Milan) and assess improvements in diagnostic accuracy, we first conducted a correlation analysis to understand variable relationships, guide feature selection, and refine our approach for the subsequent logistic regression. The correlation analysis confirmed a positive association between plasma Fibulin-3 levels and fibrosis stage (r = 0.40, p < 0.0001), while no significant correlations were observed with steatosis grade (r = 0.12, p = 0.07), lobular inflammation (r = 0.19, p = 0.05), or hepatocellular ballooning (r = 0.08, p = 0.55) (**Table S3**).

We then conducted a logistic regression using hierarchical forward selection and a switching mode, which selected the best parameter combinations from 18 continuous clinic-biochemical variables (age, weight, height, BMI, excess weight, AST, ALT, GGT, fasting glucose, triglycerides, HDL-cholesterol, platelets, Albumin, ALT/AST, APRI, FIB-4, FORNS, HSI, among others). The resulting diagnostic algorithm increased the AUROC from 0.78 for plasma Fibulin-3 alone to 0.87 when combined with platelets, GGT, and HSI (**Figure 5A**). Applying the equation of the algorithm (-4.34 + 0.11 X plasma Fibulin-3 - 0.01 X PLT + 0.11 X HSI + 0.02 X GGT) correctly classified 78.9% of the selected samples in the MO combined cohort (**Figure 5B**).

**Figure 5.**
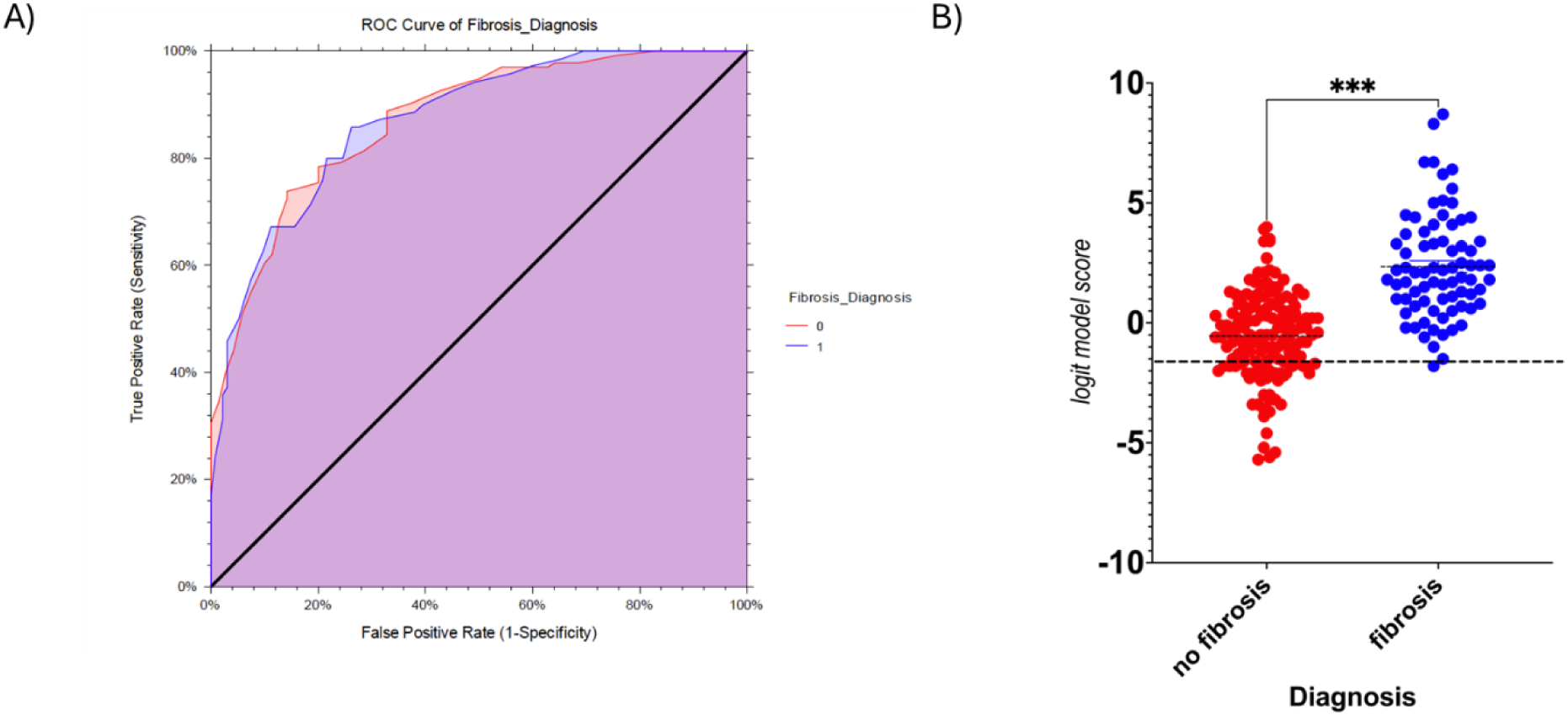
Logistic Regression Analysis. (A) AUROC for combined biomarkers B) Comparison of logit model score between FBLN-3 plasma with or without fibrosis. The optimal threshold is represented by a dashed line.

## 4. Discussion

Despite the continuously rising prevalence of MASLD, particularly of its progressive form, MASH, most affected individuals remain clinically silent or present only with vague symptoms such as fatigue or upper abdominal discomfort (22). Current diagnostic solutions face a myriad of limitations. Routine blood-based biochemical markers, such as aminotransferases, γ-glutamyl transpeptidase, and platelet counts, are non-specific and may remain within normal ranges even in progressive disease in certain diagnoses (23). While imaging techniques such as abdominal ultrasound and transient elastography are commonly used to detect steatosis and fibrosis, their sensitivity is compromised in individuals with high body mass index or in early-stage disease (24,25). In addition the limitations of non-invasive scoring and elastography increase in patients with severe obesity. More advanced techniques, such as magnetic resonance imaging, proton density fat fraction, and magnetic resonance elastography, provide greater accuracy in measuring liver fat and fibrosis; however, they are costly and mostly limited to academic centres (26–29). Furthermore, the long-standing diagnostic gold standard, liver biopsy, is invasive, carries procedural risks, and is unsuitable for routine use or population-wide screening. Given these challenges, there remains an urgent clinical need for novel, non-invasive biomarkers capable of accurately distinguishing early-stage fibrosis from moderate or advanced fibrosis in MASLD (3,19).

To address this need, our study employed an integrative multi-step strategy, combining *in-silico* analysis with experimental determination in plasma and liver samples. This approach successfully identified Fibulin-3 (*EFEMP1* gene product) as a promising candidate due to its differential expression in the liver and blood circulation. Fibulin-3 is a secreted glycoprotein, incorporated in elastic fiber networks and crucial for maintaining ECM integrity and regulating cell-matrix interactions (30,31).

Our findings align strongly with recent studies that highlighted Fibulin-3’s role in liver disease, noting its upregulation in fibrotic liver tissues and its association with hepatic stellate cell activation (32,33). Zheng et al. demonstrated that EFEMP1 is part of a liver gene network that is inhibited in healthy subjects but activated in MASL and MASH groups (34). *EFEMP1* silencing in LX-2 cells has been shown to disrupt fibronectin organization, impair focal adhesion signalling, and reduce cell migration, underscoring its role in ECM assembly(35). Interestingly, Moylan et al. have identified a 64-gene molecular signature that includes *EFEMP1*, significantly upregulated in severe MASLD compared to mild MASLD (30). Beyond the liver, dysregulated fibulin expression is a common feature in diverse pathological contexts, including fibrosis and cancer, where fibulins are enriched in sites of epithelial-mesenchymal transition (30,36). Consistent with these observations, we found a progressive increase in circulating Fibulin-3 levels across fibrosis stages in our morbidly obese cohort, supporting its potential as a measurable marker of ECM remodelling and fibrosis progression in MASH.

The diagnostic performance of circulating Fibulin-3 was optimal for distinguishing between moderate/advanced fibrosis and minimal fibrosis in our study (total samples, n=225). This finding warrants additional experimental validation in large, multi-center cohort studies. Our results resonate with previous research suggesting that other circulating ECM fragments (e.g., PRO-C3, TIMP-1, osteopontin) reflect fibrogenic activity. However, Fibulin-3 is notable for its dual role as both a structural matrix protein and a signalling regulator of fibrosis-related pathways, including TGF-β/Smad signalling (32–34). At the gene expression level, multiple transcriptome studies have shown increased *EFEMP1* expression in fibrotic hepatic tissues (35–38), and Giraudi et al. independently identified EFEMP1 as one of the top hub genes associated with liver fibrosis (10). Furthermore, at the circulating level, Sakane et al. identified Fibulin-3 as significantly enriched in extracellular vesicles from patients with advanced fibrosis, correlating with an increased risk of liver-related events during follow-up (37). In line with that work, our study emphasizes the diagnostic value of Fibulin-3 for moderate/advanced fibrosis stages in a cross-sectional MASLD cohort, particularly in high-risk morbidly obese individuals.

Analysis of the diagnostic performance revealed that plasma Fibulin-3 had superior AUROC values compared to standard non-invasive fibrosis scores, such as FIB-4, APRI, and NFS, at least in morbidly obese cohorts. Its AUROC was also comparable with other MASH-fibrosis biomarkers like PRO-C3 (11). Crucially, when integrated with other biochemical parameters (GGT, HSI, PLT), the Fibulin-3 marker vastly improved diagnostic accuracy, demonstrating its strong potential in composite algorithms for non-invasive fibrosis staging. This is a critical finding, given that a recent study from the LITMUS project showed no single biomarker or multi-marker score significantly met the predefined AUROC threshold of 0.80 for clinically significant fibrosis. Our initial findings in the CPF cohort also suggest that plasma Fibulin-3 can differentiate between minimal fibrosis (F0-F1) and moderate/advanced fibrosis (F2-F3) in the general overweight and obese population, which strengthens our hypothesis about its potential as a broad diagnostic marker for fibrosis in MASLD. Some limitations of our study must be mentioned. The cohort sample size was relatively small, particularly with a low number of participants with advanced fibrosis or cirrhosis (4%). We relied solely on ELISA for protein quantification, and we did not explore variables such as the source (e.g., EV vs. total circulating protein) or the influence of different matrix types. Nevertheless, our findings demonstrate that EFEMP1/Fibulin-3 is a robust and reliable biomarker for diagnosing fibrosis in morbidly obese cohorts. The use of two independent cohorts, one histologically diagnosed and one with fibrosis estimated by elastography, provides initial validation for its utility, and the extension of this study in large multicentre cohorts is foreseen.

## 5. Conclusion

We developed a new *in-silico* strategy that combines large-omics data, such as transcriptome and proteome datasets, to identify novel biomarkers capable of diagnosing fibrosis in the high-risk morbidly obese population. Plasma Fibulin-3 was identified as a strong candidate whose consistent upregulation in fibrotic liver tissue and measurable increase in circulation support its use as both a standalone marker and part of a multi-marker diagnostic index. When used in conjunction with other non-invasive markers, plasma Fibulin-3 can significantly enhance the diagnostic accuracy for detecting and staging fibrosis in MASLD.

## Data Availability

All data produced in the present work are contained in the manuscript

## Acknowledgements

The authors would like to thank all the participants in the morbidly obese cohort. We also appreciate our colleagues at Fondazione Italiana Fegato, as well as our collaborators, clinicians, and nurses from Cattinara Hospital, Centro Patologie di Trieste, and Policlinico di Milano for their kind assistance and collaboration. AI-assisted technologies, ChatGPT-5 and Grammarly Pro www.grammarly.com (v1.2.192.1748), were used to enhance the language and readability of the manuscript.

## Ethics approval and consent to participate

All enrolled patients provided written consent, and the local Ethics Committee approved the studies under these protocols: for the Trieste bariatric cohort, study protocol No. 22979 (Comitato Etico Regionale Unico, FVG, SSN, Italy). This study is also registered at ClinicalTrials.gov under ID NCT06098417; for the Milan bariatric cohort, the study protocol was approved with code CE 401 (02/28/2019) by the Ethical Committees of Fondazione IRCCS Cà Granda, Milan. For the CPF obese cohort, the study was approved according to CEUR, FVG, under protocol 17385, and with the internal healthcare code 261_2023H.

## Consent for publication

All authors have reviewed the final version of the manuscript and approved it for submission.

### Abbreviations

ALT: alanine aminotransferase;
AST: aspartate aminotransferase;
AUROC: Area Under the Receiver Operating Curve;
BCA: bicinchoninic assay;
BMI: body mass index;
FFA: free fatty acids;
GGT: gamma-glutamyl transferase;
HDL: high-density cholesterol;
MASLD: metabolic dysfunction-associated steatotic liver disease;
MASH: metabolic-associated steatohepatitis;
MO: morbidly obese;
NAFLD: nonalcoholic fatty liver disease;
T2DM: type 2 diabetes mellitus.

