## Supplementary information for "Fibulin-3 as a reliable biomarker of fibrosis in obese subjects with Metabolically Dysfunction Associated Steatotic Liver Disease"

**Materials and Methods**

*Clinical assessment*

At the baseline visit, anthropometric parameters, including age, sex, body weight, height, BMI calculation, and waist circumference, were recorded. After overnight fasting, blood samples were collected before surgery to determine glucose, liver biochemistry (AST, ALT, GGT), albumin, platelets, lipid profile (TG, T-Chol, HDL), among others. Diabetes was diagnosed according to the ESC-EASD guidelines (21). Blood-based tests of liver fibrosis, such as FIB-4, APRI, FORNS, and NFS were calculated as described (5–7).

*Quantitative PCR*

Total RNA was extracted from cell culture lysate and homogenized liver tissues using Tri-reagent kit (Sigma-Aldrich, MO, USA). cDNA synthesis was performed with High-Capacity cDNA Reverse Transcription Kit (Applied Biosystems, Waltham, MA, USA). The specific primer pairs for the selected genes were designed for use in experimental mRNA expression analysis by quantitative reverse transcription polymerase chain reaction (RT-qPCR) Quantitative PCR was performed in CFX Connect Real-Time PCR Detection System (Bio-Rad, Hercules, CA USA) in a specific reaction volume containing 25 ng of cDNA, 1X iQ SYBR Green Supermix, and primer pairs. Primer sequences for both human genes are specified in Table 5. Relative gene expression was calculated using Pfaffl modification of the ΔΔCt method, normalizing all measurements with housekeeping genes RPLP0 and TUBB1A1 and considering the previously determined efficiencies for each gene of interests. Expression data were represented as mean ± standard deviation. The sequence of used primers is in Table S4.

*Western Blot Analysis*

Liver tissue samples were homogenized in HNTG lysis buffer (50mM HEPES, 150mM NaCl, 10% glycerol, 1% Triton X-100, pH 7.5). Protein quantification of liver homogenates was done using the Bicinchoninic acid (BCA) method, and the samples were stored at -80°C until further use.

Liver homogenates (100 g) were solubilized in 5X Laemmli Buffer with 10% ß-mercaptoethanol and denatured at 95°C for 5 minutes. Each sample were loaded to their respective wells onto the stacking gel of 12% polyacrylamide sodium dodecyl sulfate-polyacrylamide gel electrophoresis (SDS-PAGE). Protein bands in the gel were transferred onto polyvinylidene difluoride (PVDF) membrane through electroblotting. Blotting was executed at 100V for 2 hours while submerged in transfer buffer composed of 25mM Tris base, 190mM glycine, and 20% methanol. PVDF membranes were incubated for 1 hour in solutions of 4% milk Tris-Buffered Saline (TBS; 20mM NaCl and 150mM)-Tween 0.1% (T-TBS) to block non-specific binding sites. The following primary antibodies were used: Fibulin-3/EFEMP1 1:250 (BT Lab, Jiaxing, Zhejiang Province, China) and the reference transferrin 1:1000 (Santa Cruz Biotech, Santa Cruz, CA, USA). Blots were incubated with anti-rabbit IgG-HRP-conjugated secondary antibody (1:1000 for Fibulin-3/EFEMP1 and 1:2000 for transferrin). Protein bands were visualized using the ECL immunoblotting detection system (GE Healthcare, Buckinghamshire, UK) and developed on a C-DiGit ® Blot Scanner (LI-COR Biosciences, NE, USA). Results were expressed as the ratio of fibulin-1 protein expression to that of a reference housekeeping protein, transferrin. Relative densitometry analyses of the immunoblots were determined using Image Studio™ Version 5.2 Acquisition software (LI-COR Biosciences, Lincoln, NE, USA).

*ELISA*

Plasma protein levels were measured by the following ELISA commercial kits: Fibulin-3 (Human FBLN3 ELISA Kit, E-EL-H1673, Elabscience), Lumican (Human Lumican ELISA Kit, E1871Hu, BT Lab), Chinitase 1-like 3 (RayBio® Human CHI3L1 ELISA Kit, ELH-CHI3L1, RayBiotech) using the manufacturers' instructions. The optimal sample dilution was established by testing serial dilutions of representative samples to ensure readings fell within the linear range of the ELISA standard curve. The final working dilution was selected to ensure precise and reliable quantification within the assay’s dynamic range.

**Abbreviations**

ALT, alanine aminotransferase; AST, aspartate aminotransferase; AUROC, Area Under the Receiver Operating Curve; BCA, bicinchoninic assay; BMI, body mass index; FFA, free fatty acids; GGT, gamma-glutamyl transferase; HDL, high-density cholesterol; MASLD, metabolic dysfunction-associated steatotic liver disease; MASH, metabolic-associated steatohepatitis; MO, morbidly obese; NAFLD, nonalcoholic fatty liver disease; T2DM, type 2 diabetes mellitus.

**Supplementary Figures**

**Figure S1.** Plasma levels for selected candidates in the discovery cohort between F0/F1 and F2-F3/F4 groups.

B) Chitinase 3-like 1 and C) Lumican. Values presented are the mean ±SD *p <0.05, **p <0.01, ***p <0.0001.

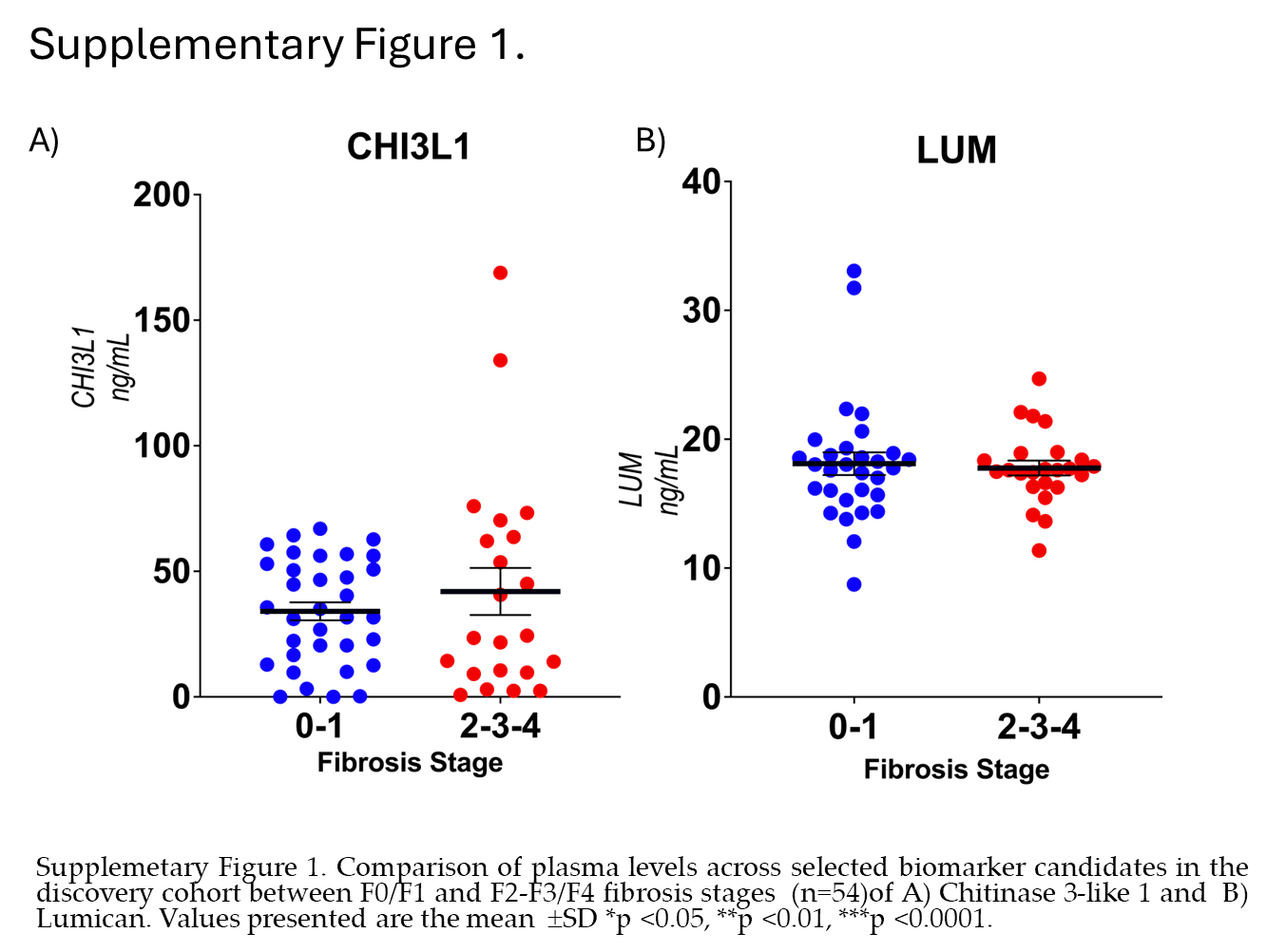

**Figure S2.**

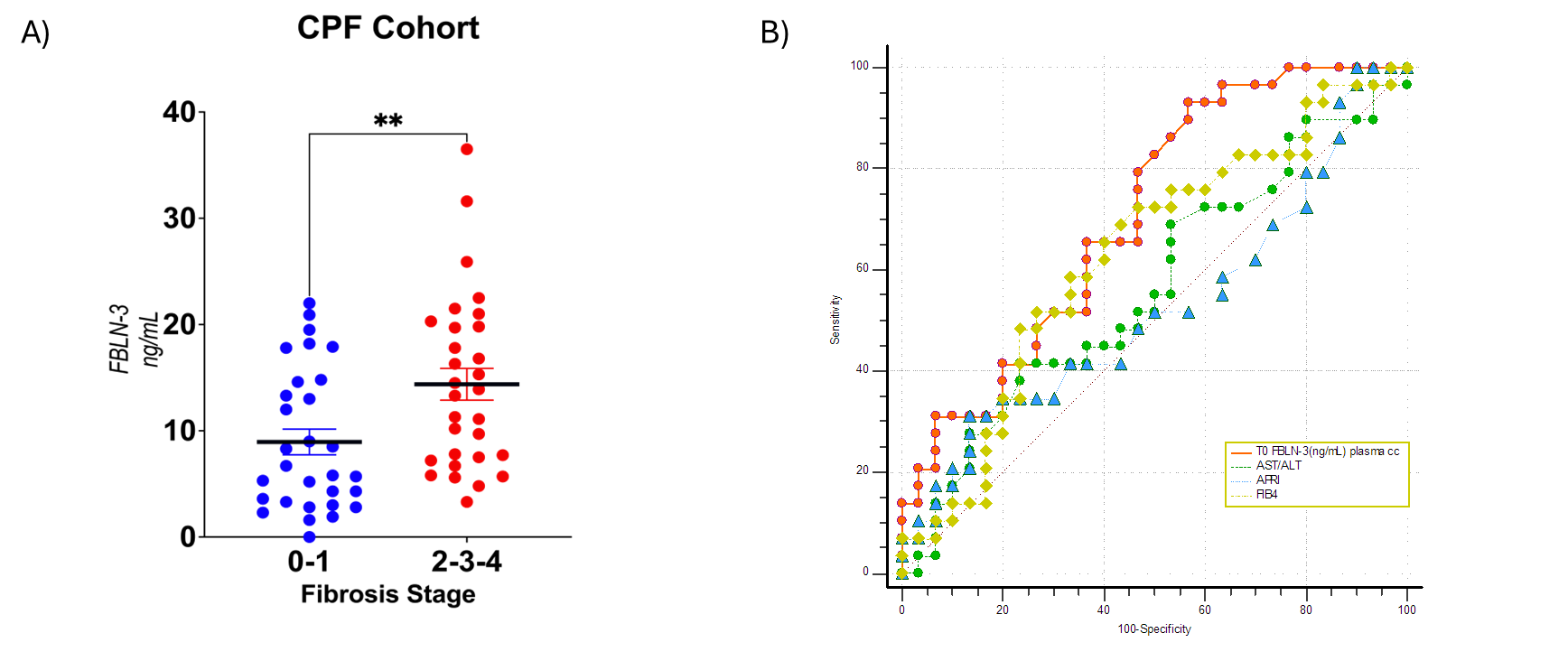

**Figure S2. *Plasma FBLN-3 and AUROC curves in the CPF cohort.*** (A) Fibulin-3 plasma levels in the CPF cohort are stratified according to fibrosis, and (B) compares AUROC curves for plasma Fibulin-3 against other blood-based scoring tests in the CPF cohort. The Chi-Square test was used for categorical variables. *** indicates significance at p < 0.001, ** at p < 0.01, and * at p < 0.05.

**Supplementary Tables**

**Table S1.** List of differentially expressed genes between F2-F3-F4 and F0-F1 fibrosis groups using 3D-RNASeq pipeline.

**Table S2.** The clinical-demographic data of the patients selected in the CPF cohort.

**Table S3.** Correlation Analyses of Plasma Fibulin-3 against several blood-based and histopathological parameters

**Table S4.** List of primers used in the study
